# Genomic and phenomic insights from an atlas of genetic effects on DNA methylation

**DOI:** 10.1101/2020.09.01.20180406

**Authors:** Josine L. Min, Gibran Hemani, Eilis Hannon, Koen F. Dekkers, Juan Castillo-Fernandez, René Luijk, Elena Carnero-Montoro, Daniel J. Lawson, Kimberley Burrows, Matthew Suderman, Andrew D. Bretherick, Tom G Richardson, Johanna Klughammer, Valentina Iotchkova, Gemma Sharp, Ahmad Al Khleifat, Aleksey Shatunov, Alfredo Iacoangeli, Wendy L McArdle, Karen M Ho, Ashish Kumar, Cilla Söderhäll, Carolina Soriano-Tárraga, Eva Giralt-Steinhauer, Nabila Kazmi, Dan Mason, Allan F McRae, David L Corcoran, Karen Sugden, Silva Kasela, Alexia Cardona, Felix R. Day, Giovanni Cugliari, Clara Viberti, Simonetta Guarrera, Michael Lerro, Richa Gupta, Sailalitha Bollepalli, Pooja Mandaviya, Yanni Zeng, Toni-Kim Clarke, Rosie M Walker, Vanessa Schmoll, Darina Czamara, Carlos Ruiz-Arenas, Faisal I Rezwan, Riccardo E Marioni, Tian Lin, Yvonne Awaloff, Marine Germain, Dylan Aïssi, Ramona Zwamborn, Kristel van Eijk, Annelot Dekker, Jenny van Dongen, Jouke-Jan Hottenga, Gonneke Willemsen, Cheng-Jian Xu, Guillermo Barturen, Francesc Català-Moll, Martin Kerick, Carol Wang, Phillip Melton, Hannah R Elliott, Jean Shin, Manon Bernard, Idil Yet, Melissa Smart, Tyler Gorrie-Stone, BIOS Consortium, Chris Shaw, Ammar Al Chalabi, Susan M Ring, Göran Pershagen, Erik Melén, Jordi Jiménez-Conde, Jaume Roquer, Debbie A Lawlor, John Wright, Nicholas G Martin, Grant W Montgomery, Terrie E Moffitt, Richie Poulton, Tõnu Esko, Lili Milani, Andres Metspalu, John R. B. Perry, Ken K. Ong, Nicholas J Wareham, Giuseppe Matullo, Carlotta Sacerdote, Avshalom Caspi, Louise Arseneault, France Gagnon, Miina Ollikainen, Jaakko Kaprio, Janine F Felix, Fernando Rivadeneira, Henning Tiemeier, Marinus H van IJzendoorn, André G Uitterlinden, Vincent WV Jaddoe, Chris Haley, Andrew M McIntosh, Kathryn L Evans, Alison Murray, Katri Räikkönen, Jari Lahti, Ellen A Nohr, Thorkild IA Sørensen, Torben Hansen, Camilla Schmidt Morgen, Elisabeth B Binder, Susanne Lucae, Juan Ramon Gonzalez, Mariona Bustamante, Jordi Sunyer, John W Holloway, Wilfried Karmaus, Hongmei Zhang, Ian J Deary, Naomi R Wray, John M Starr, Marian Beekman, Diana van Heemst, P Eline Slagboom, Pierre-Emmanuel Morange, David-Alexandre Trégouët, Jan H. Veldink, Gareth E Davies, Eco JC de Geus, Dorret I Boomsma, Judith M Vonk, Bert Brunekreef, Gerard H. Koppelman, Marta E Alarcón-Riquelme, Rae-Chi Huang, Craig Pennell, Joyce van Meurs, M. Arfan Ikram, Alun D Hughes, Therese Tillin, Nish Chaturvedi, Zdenka Pausova, Tomas Paus, Timothy D Spector, Meena Kumari, Leonard C Schalkwyk, Peter M Visscher, George Davey Smith, Christoph Bock, Tom R Gaunt, Jordana T Bell, Bastiaan T. Heijmans, Jonathan Mill, Caroline L Relton

**Author notes:** These authors contributed equally to this research (Josine L. Min, Gibran Hemani). **Corresponding author:** Josine L Min.

## Abstract

Characterizing genetic influences on DNA methylation (DNAm) provides an opportunity to understand mechanisms underpinning gene regulation and disease. Here we describe results of DNA methylation-quantitative trait loci (mQTL) analyses on 32,851 participants, identifying genetic variants associated with DNAm at 420,509 DNAm sites in blood. We present a database of >270,000 independent mQTL of which 8.5% comprise long-range (*trans*) associations. Identified mQTL associations explain 15-17% of the additive genetic variance of DNAm. We reveal that the genetic architecture of DNAm levels is highly polygenic and DNAm exhibits signatures of negative and positive natural selection. Using shared genetic control between distal DNAm sites we construct networks, identifying 405 discrete genomic communities enriched for genomic annotations and complex traits. Shared genetic factors are associated with both blood DNAm levels and complex diseases but in most cases these associations do not reflect causal relationships from DNAm to trait or vice versa indicating a more complex genotype-phenotype map than has previously been hypothesised.

## Main

The role of inter-individual variation in DNA methylation (DNAm) on disease mechanisms is not yet well characterised. It has, however, been hypothesised to serve as a viable biomarker for risk stratification, early disease detection and the prediction of disease prognosis and progression^1^. Because genetic influences on DNAm have been shown to be widespread^2,3,4^, a powerful avenue into researching the functional consequences of changes in DNAm levels is to map genetic differences associated with population-level variation, identifying DNA methylation quantitative trait loci, (mQTL) that include both local (*cis* mQTL) and distal (*trans* mQTL) effects. We can harness mQTL as natural experiments, allowing us to observe randomly perturbed DNAm levels in a manner that is not confounded with environmental factors^5,6^. In this regard, mapping even very small genetic effects on DNAm is valuable for gaining power to evaluate whether its variation has a substantial causal role in disease and other biological processes.

To date, only a small fraction of the total genetic variation estimated to influence DNAm across the genome has been identified, primarily restricted to *cis* mQTL^7^. Yet the majority of genetic effects are likely to act in *trans* (defined as more than 1Mb from the DNAm site) and have small effect sizes^5,7-9^. As with classical complex traits^10^, much larger sample sizes are required to map associations involving small genetic effects in order to permit greater understanding of the genetic architecture and the biological processes underlying DNAm^7^. To this end, we established the Genetics of DNA Methylation Consortium (GoDMC), an international collaboration of human epidemiological studies that comprises >30,000 study participants with genetic, phenotypic and DNAm data.

Importantly, the unrivalled sample size and coverage of our study enables us to identify a large number of *cis* and *trans* mQTL to gain biological insights that were previously impossible. First, we use this extensive resource to uncover the genetic architecture of DNAm and to study natural selection pressures. Second, we learn about how *cis-* and *trans-*acting variants and DNAm sites interact through the development of new network approaches. Third, we interrogate the potential role of DNAm in disease mechanisms by exhaustively mapping the causal relationships of DNAm with 116 complex traits and diseases in a bi-directional manner. A database of our results is available as a resource to the community at http://mqtldb.godmc.org.uk/.

### Genetic variants influence 45% of tested DNAm sites

In order to map genetic influences on DNAm, we established an analysis workflow that enabled standardized meta-analysis and data integration across 36 population-based and disease datasets with genotype and DNAm data. Using a two-phase discovery study design, we analyzed ~10 million genotypes imputed to the 1000 Genomes reference panel^11^ and 420,509 DNAm sites measured by Infinium HumanMethylation BeadChips in whole blood derived from 27,750 European participants (**Figures 1A** and **S1-S5**, **Table S1-S2, Supplementary Note 1, Supplementary Information**).

**Figure 1:**
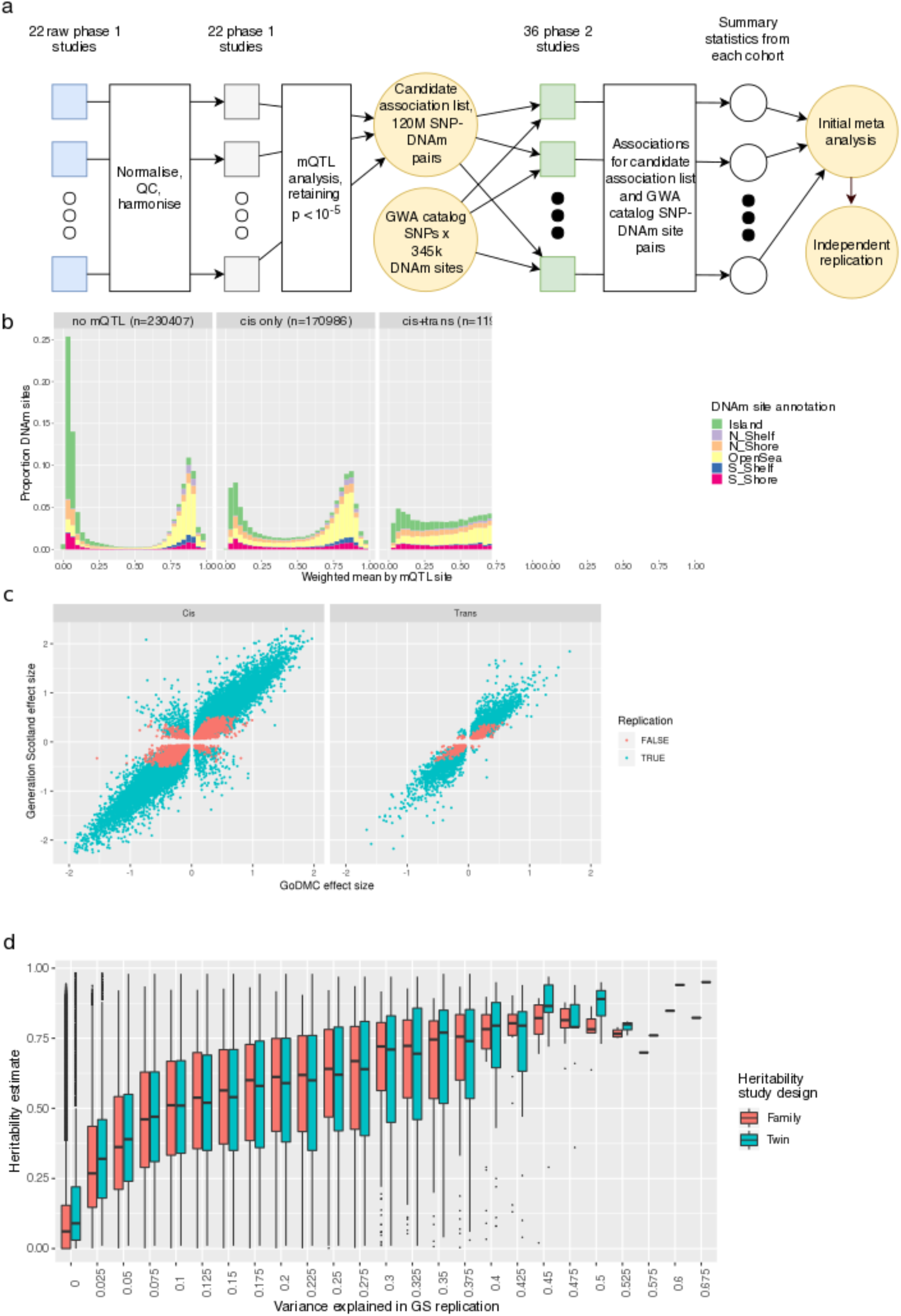
Discovery and replication of mQTL. **a) Study Design**. In the first phase, 22 cohorts performed a complete mQTL analysis of up to 480,000 sites against up to 12 million variants; retaining their results for p<1e-5. In the second phase, 120 million SNP-DNAm site pairs selected from the first phase, and GWA catalog SNPs against 345k DNAm sites, were tested in 36 studies (including 20 phase 1 studies) and metaanalysed. **b) Distributions of the weighted mean of DNAm across 36 cohorts for *cis only, cis+trans* and *trans only* sites**. Plots are coloured with respect to the genomic annotation. *Cis only* sites showed a bimodal distribution of DNAm. *Cis+trans* sites showed intermediate levels of DNAm. *Trans only* sites showed low levels of DNAm. **c) Discovery and replication effect size estimates** between GoDMC (n=27,750) and Generation Scotland (n=5,101) for 169,656 mQTL associations. The regression coefficient is 1.13 (se=0.0007). **d) Relationship between DNAm site heritability estimates and DNAm variance explained in Generation Scotland**. The regression coefficient for the twin family study was 3.16 (se=0.008) and for the twin study 2.91 (se=0.008) across 403,353 DNAm sites. The variance explained for DNAm sites with missing r^2^ (n=277,428) and/or h^2^=0 (Twin family: n=80,726 Twin: 34,537) were set to 0.

Using linkage disequilibrium (LD) clumping, we identified 248,607 independent *cis*-mQTL associations (p < 1e-8, < 1Mb from the DNAm site, **Figure S4**) with a median distance between single nucleotide polymorphisms (SNP) and DNAm sites of 36kb (IQR=118 kb, **Figure S3A**). We found 23,117 independent *trans* mQTL associations (using a conservative threshold of p < 1e-14^7^, **Figure S4, Supplementary Information**). These mQTL involved 190,102 DNAm sites, representing 45.2% of all those tested (**Figure 1B**) which is a 1.9x increase of sites with a *cis* association (p<1e-8) and 10x increase of sites with a *trans* association (p<1e-14) over a previous study whose sample size was 7x smaller^8^. As expected, mQTL effect sizes for each DNAm site (the maximum absolute additive change in DNAm level measured in standard deviation (SD) per allele) were lower for sites with a *trans* association (as compared to sites with a *cis* association (per allele SD change = -0.02 (s.e.=0.002, p=2.1e-14, **Figure S6**). The differential improvement in yield between *cis* and *trans* associations is revealing in terms of the genetic architecture – relatively small sample sizes are sufficient to uncover the majority of large *cis* effects, whereas much larger sample sizes are required to identify the polygenic *trans* component.

The majority of *trans* associations (80%) were inter-chromosomal. Of the intra-chromosomal *trans* associations, 34% were >5 Mb from the DNAm site, **Figure S7**). We then compared the rate of inter-chromosomal *trans* associations to the rate of intra-chromosomal *trans* associations (excluding chromosome 6) and found a substantially lower number of inter-chromosomal *trans* associations per 5 Mb region (1.59) than intra-chromosomal associations (>1 Mb: 7.95; >6 Mb 4.81).

Next, using conditional analysis^12^ we explored the potential for multiple independent SNPs operating within the locus of each mQTL, identifying 758,130 putative independent variants. Each DNAm site, for which a mQTL in *cis* had been detected, had a median of 2 independent variants (IQR=4 variants, **Figure S8**). For all subsequent analyses, we used index SNPs from clumping procedures to be conservative and unbiased due to the non-independence of genetic variants.

We sought to replicate these mQTL using the Generation Scotland (GS) cohort (n = 5,101) for which mQTL results were previously generated using an independent analysis pipeline (**Supplementary Information, Supplementary Note 1**). Data were available to allow us to test for replication of 188,017 of our discovery mQTL (137,709 sites) and we found a very strong correlation of effect sizes for both *cis* and *trans* effects (r=0.97, n=155,191 and 0.96, n=14,465 at p<1e-3, respectively; **Figure 1C**); 99.6% of the associations had a consistent sign (further discussion in **Supplementary Information**). At an approximate Bonferroni corrected threshold of 0.05/188,017, 142,727 of the discovery mQTL replicated in the GS cohort (76%); the replication rate for *cis* and *trans* mQTL were 76% and 79%, respectively. To evaluate whether our replication rate was in line with expectations given the smaller replication sample size, we estimated that under the assumption that the discovery mQTL are true positives 171,824 mQTL would be expected to replicate at a nominal threshold of 1e-3. In very close agreement we found that the actual number of mQTL replicating at this level was 169,656, indicating that the majority of our discovery mQTL are likely to be true positives (**Table S3, Supplementary Information**). Our findings support that there is little between-study heterogeneity in our analysis and that genetic effects on DNAm are highly stable across cohorts (**Figure S2, Table S2**).

Overall the variance explained by replicated genetic effects was small. For 99% of the associations in *cis* and *trans*, mQTL explained less than 21% and 16% of the DNAm variation respectively (**Figure S9**). Aggregating across all 420,509 tested DNAm sites, our replicated mQTL associations explain 1.3% of the total assayed DNAm variation, 8% of this being due to *trans-*associations. Restricting to sites that have at least one *cis-*effect or *trans-*effect, however, we explain 4.2% and 2.5% of the DNAm variance, respectively.

We then investigated how much of the heritability of variable DNAm can be explained by mQTL associations in GoDMC using family-based heritability studies of DNAm^2,3^. We found a strong positive relationship between variance explained by replication mQTL estimates (127,680 sites in GS) and heritability for both studies (family: r=0.41 across, 121,582 available sites; twin: r=0.37 across 118,955 available sites) (**Figure 1D, Table S4**). The mQTL that we identified explain 15%-17% of the additive genetic variance of DNAm (**Figure S10**). Finally, there were strong positive relationships between the heritability of DNAm levels at a DNAm site and the number of independent mQTL (**Figure S11**), heritability and effect size (**Figure S12**), variance explained and the number of independent mQTL (**Figure S13**) and variance explained and distribution of DNAm levels (**Figure S14**). Overall, our results support a mixed genetic architecture of polygenic genome-wide effects and larger *cis* effects.

### *Cis* and *trans* mQTL operate through distinct mechanisms

DNAm is usually associated with gene repression, with loss of DNAm reflecting enhancer or gene activation^13-15^. We analysed how interindividual DNAm changes are associated to genetic variation in a context way which has so far mainly focused on *cis* mQTL^7,8,16,17,19^. The statistical power of the mQTL analysis allowed us to identify SNPs only associated with DNAm in *cis* (n=157,095, 69.9%), only associated with DNAm in *trans* (n=794, 0.35%), or associated with DNAm in both *cis* and *trans* (n=66,759, 29.7%). Similarly, of the 190,102 DNAm sites influenced by a SNP, 170,986 DNAm sites (89.9%) were *cis-only*, 11,902 DNAm sites (6.3%) were *cis+trans*, and 7,214 DNAm sites (3.8%) were *trans-only*. This categorisation allowed us to infer biological properties of *trans-*features that were not due to their *cis-*effects.

Here, we first compared the distribution of DNAm levels (weighted mean DNAm level across 36 studies (defined as low (<20%), intermediate (20%-80%) or high (>80%) between the *cis* and *trans* DNAm sites (**Figure 1B**). We then performed enrichment analyses on the mQTL SNPs and DNAm sites using 25 combinatorial chromatin states from 127 cell types (including 27 blood cell types)^18^ and gene annotations (**Figure 2A, S15-S18, Tables S5-S8**). Consistent with previous studies^7,8,19^, we found that *cis only* sites are represented in high (32%), low (28%) and intermediate (40%) DNAm levels and these sites are mainly enriched for enhancer chromatin states (mean OR=1.37), CpG islands (OR=1.25) and shores (OR=1.26).

**Figure 2:**
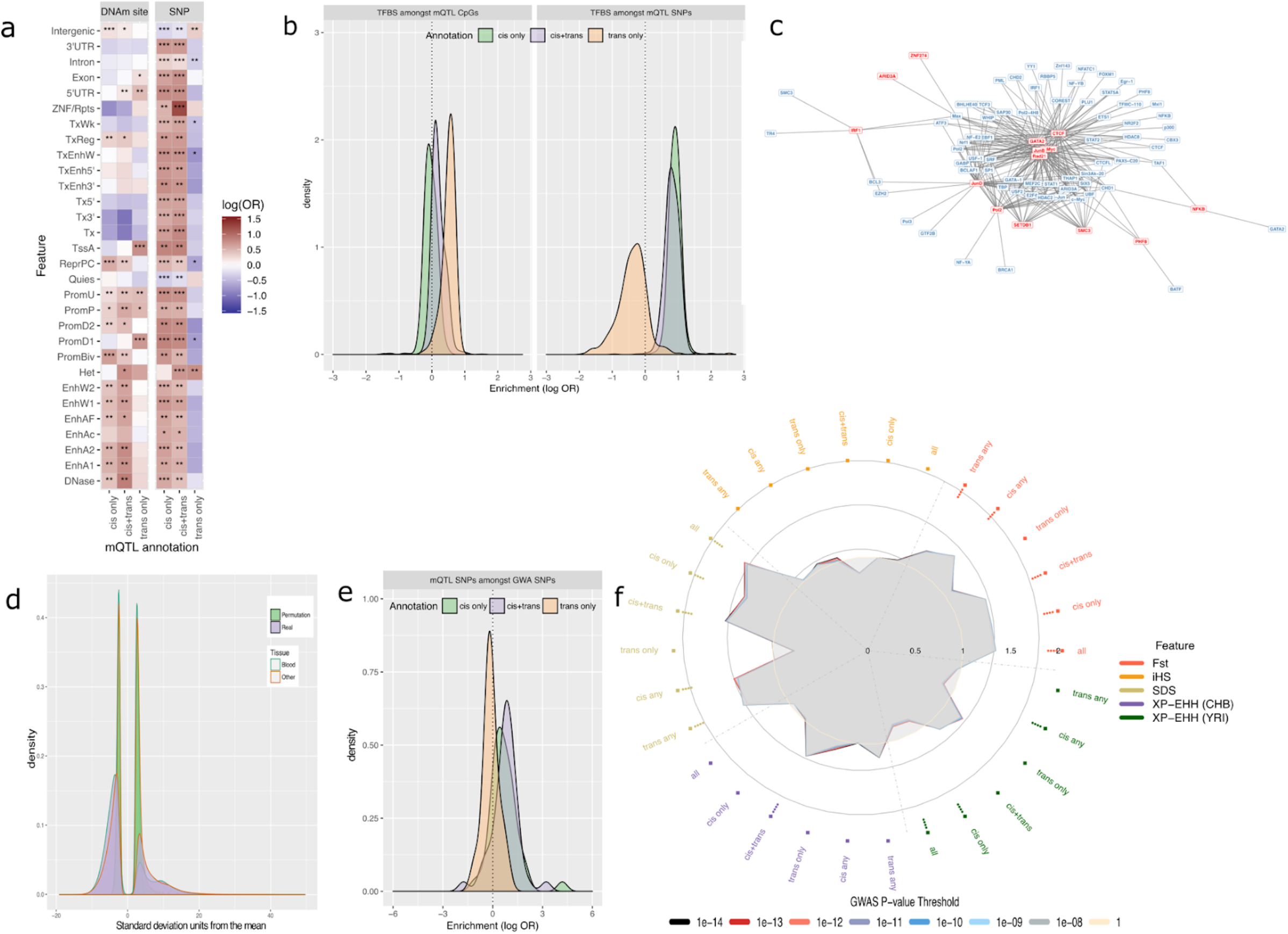
*Cis* and *trans* mQTL operate through distinct mechanisms. **a) Distributions of enrichments for chromatin states and gene annotations among mQTL sites and SNPs**. The heatmap represents the distribution of odds ratios for *cis only, trans only*, or *cis+trans* sites and SNPs. Significance has been categorised as: *=FDR<0.001;**=FDR<1e- 10;***=FDR<1e-50 **b) Distributions of enrichment for occupancy of TFBS among mQTL sites and SNPs**. Each density curve represents the distribution of odds ratios for *cis only, trans only*, or *cis+trans* sites (left) and SNPs (right). **c) A bipartite graph of the two-dimensional enrichment for *trans-*mQTL**. SNPs annotations (blue) with p_emp_ < 0.01 after multiple testing correction co-occur with particular site annotations (red). **d) Distribution of two-dimensional enrichment values of *trans-*mQTL**. There was substantial departure from the null in the real dataset for all tissues indicating that the TFBS of a site depended on the TFBS of the SNP that influenced it. **e) Distributions of enrichment of mQTL among 42 complex traits and diseases**. Each density curve represents the distribution of odds ratios for *cis only, trans only*, or *cis+trans* SNPs. **f) Enrichment of selection signals among mQTL SNPs**. Radial lines show odds ratios for the different selection metrics (F_st_, SDS, iHS, XPEHH (CEU vs CHB) and XPEHH (CEU vs YRI) by site annotation (*cis any, cis only, cis+trans, trans only, trans any*). Dots in the inner ring of the outer circle denote enrichment (if present) at thresholds p<1e-11 (outermost) to p<1e-14 (innermost).

For *cis+trans* sites, we found that the majority of these sites (66%) have intermediate DNAm levels. By replicating this finding in two isolated white-blood-cell subsets, we showed that this is due to cell-to-cell variability^18,20^ and not explained by between cell type differences (**Figure S19**). In line with the observation that intermediate levels of DNAm are found at distal regulatory sequences^21,24^, these sites were enriched for enhancer (mean OR=1.65) and promoter states (mean OR=1.41). However for *trans only* sites, we found a pattern of low DNAm (for 55% of sites) and enrichments for promoter states (mean OR=1.39) especially TssA promoter state (mean OR=2.03). We demonstrated that these inferences about *cis* and *trans* enrichments were not sensitive to the definition of *trans* associations, by showing that the patterns were consistent if we restricted to only inter-chromosomal associations (**Supplemental Information, Figure S20**).

We continued by analysing the differences in properties between SNPs that have local versus long-range DNAm influences. We found that *cis only* and *cis+trans* SNPs were enriched for active chromatin states and genic regions whereas *trans only* SNPs were enriched for intergenic regions and the heterochromatin state (**Figure 2A, S17-S18, Tables S7-S8**). This implies that the enrichments for the *cis+trans* SNPs were driven by their *cis* signals. Overall, these results highlight that a complex relationship between molecular features is underlying the mQTL categories and the biological contexts are substantially different between *cis* and *trans* features.

We found that these inferences were often shared across other tissues. For example, DNAm sites with low or intermediate DNAm levels have similar DNAm distributions in 12 tissues (**Figure S21-23**). However, while SNP and DNAm site enrichments were typically present in multiple tissues, enrichments were stronger in blood datasets for the enhancer states (SNP: difference in mean OR=0.055, p=0.038; sites: difference in mean OR=0.21, p < 2e-16) and DNAse state (SNP: difference in mean OR=0.13, p=0.004; sites: difference in mean OR=0.41 p=9.65e-16) indicating some level of tissue specificity for mQTL in these regions (**Figure S15, S17, S24**).

To investigate the question of tissue specificity further, we compared the correlation of effect estimates of *cis* and *trans* mQTL in blood against adipose tissue (n=603)^22^ and brain (n=170)^9^ (**Supplementary Information, Table S9**). Generally, the between tissue effect correlations were high, in line with a recent comparison of *cis-*mQTL effects between brain and blood^23^. However, we found that the highest correlations were for associations involving *trans-only* sites (Adipose r_b_=0.92 (se =0.004); Brain r_b_=0.88 (se=0.009)) despite having on average smaller effect sizes than *cis only* associations, implying that they are *less* tissue specific than *cis* effects (Adipose r_b_=0.73 (se =0.002); Brain r_b_=0.59 (se=0.004)). Stratifying the mQTL categories to low, intermediate and high DNAm, showed that the brain-blood correlations are the lowest for intermediate DNAm categories and adipose-blood correlations are lowest for high DNAm categories, which may suggest cellular heterogeneity for high DNAm levels (**Table S9**). These results show the value of large sample sizes in blood to detect *trans* mQTL regardless of the tissue.

### *Trans* mQTL SNPs and DNAm exhibit patterned TF binding

Binding of transcription factor (TF) to distal regulatory elements correlates with low DNAm levels and regulates gene activity^24,25^. To gain insights into how SNPs induce long-range DNAm changes, we mapped enrichments for DNAm sites and SNPs across binding sites for 171 TFs in 27 cell types^26,27^. We found strong enrichments for the majority of TFs amongst DNAm sites with a *trans* association (*cis+trans:* 55%; *trans only:* 80%; *cis only:* 18%) and amongst *cis-acting* SNPs (*cis only:* 96%, *cis+trans:* 91%, *trans only:* 1%) (**Figures 2B, S25, S26**). Sites that overlap TFBS were relatively hypomethylated independent of tissue (weighted mean DNAm levels = 21% vs 52%, p<2.2e-16) (**Figure S27**) and we found that generally the TFBS enrichments were not tissue specific (**Table S10-11, Figure S25-26**). Overall, these enrichments illustrate multiple mechanisms at play: that marginally the *trans* associations are not arising due to direct genetic influences on TF activity, whereas *cis* associations could be. It therefore raises the question, what is the relationship between the TFBS occupancy of a DNAm site and a SNP in a *trans-*mQTL?

The enrichment analyses above have focused on analysing mQTL SNPs and mQTL sites marginally. However, a mQTL has a pair of TFBS annotations^26^, one for the SNP and one for the DNAm site, and they can be analysed for joint pairwise enrichment. Using a novel approach (two-dimensional functional enrichment, **Figure S28**), we evaluated if the annotation pairs amongst 18,584 inter-chromosomal *trans-*mQTL were associated to TF binding in a non-random pattern (**Supplementary Information**). We found that across the *trans-*mQTL there was substantial non-random pairing of TFBS annotations for DNAm sites and SNPs. We found that 6.1% (22,962 of 378,225) of possible pairwise combinations of SNP-DNAm site annotations were more over- or under-represented than expected by chance after strict multiple testing correction (**Supplementary Information, Table S12, Figure 2C-D**).

After accounting for abundance and other characteristics, the strongest pairwise enrichments involved sites close to TFBS for proteins in the cohesin complex, for example CTCF, SMC3 and RAD21, as well as TFs such as GATA2 related to cohesin^28^. Bipartite analysis showed that these clustered due to being related to similar sets of SNP annotations (**Figure 2C**). Other clusters were also found, for example, sites close to TFBS for interferon regulatory factor 1 (*IRF1*), a gene for which *trans-acting* regulatory networks have been previously reported^29^, were more likely to be influenced by SNPs near TFBS for EZH2, SMC3, ATF3, BCL3, TR4 and MAX. The relationship between IRF1 and these other proteins has been documented previously^30–32^. For example EZH2 mediates the silencing of IRF1^33^; BCL3 and IRF1 are co-down-regulated during inflammation^30^; and ATF3 is a negative regulator of cytokines which themselves induce IRF1^31,32^.

To further investigate mechanisms by which inter-chromosomal *trans* mQTL (n=18,584) can arise, we compared their locations to known regions of chromosomal interactions (genomic regions that have been shown to spatially colocalise within the cell^34^). We found 1175 overlaps for 637 SNP-DNAm site pairs (3.4%) where the LD region of the mQTL SNP and the corresponding site overlapped with any interacting regions (525 SNPs, 602 sites) as compared to a mean of 473 SNP-DNAm site pairs in 1000 permuted datasets (OR=1.36, p_Fisher_=6.5e-7, p_empirical_<1e-3) (**Figure S29**). To summarise, the order of operation between transcription factor (TF) binding and demethylation requires further investigation^8,25,16^, but these results indicate that for a small proportion of interchromosomal *trans* mQTL the spatial distance *in vivo* is likely to be small.

### Communities of DNAm sites are identified by shared *trans-*genetic effects

*trans-mQTL* provide an opportunity to infer how distal genomic regions are functionally related, but the polygenic nature of DNAm variation could lead to apparent shared genetic effects that arise from distinct causal variants rather than shared genetic factors. We observed that there were 1,728,873 instances where a SNP acting in *trans* also influenced a *cis* DNAm site (before LD pruning). Genetic colocalization analysis indicated that 278,051 of these instances were due to the *cis* and *trans* sites sharing a genetic factor, representing 3,573 independent *cis-trans* genomic region pairs, of which 3,270 were inter-chromosomal (**Table S13**). These pairs consisted of 1,755 independent SNPs and 5,109 independent DNAm sites across the genome, indicating that some sites with *cis* associations shared genetic factors with multiple sites with *trans* associations. From the *cis-trans* pairs we constructed a network linking these genomic regions which elucidated 405 “communities” of genomic regions that were substantially connected (**Supplementary Information**). Fifty-six of these communities comprised 10 or more sites, and the largest community comprised 253 sites (**Figure 3A**).

**Figure 3:**
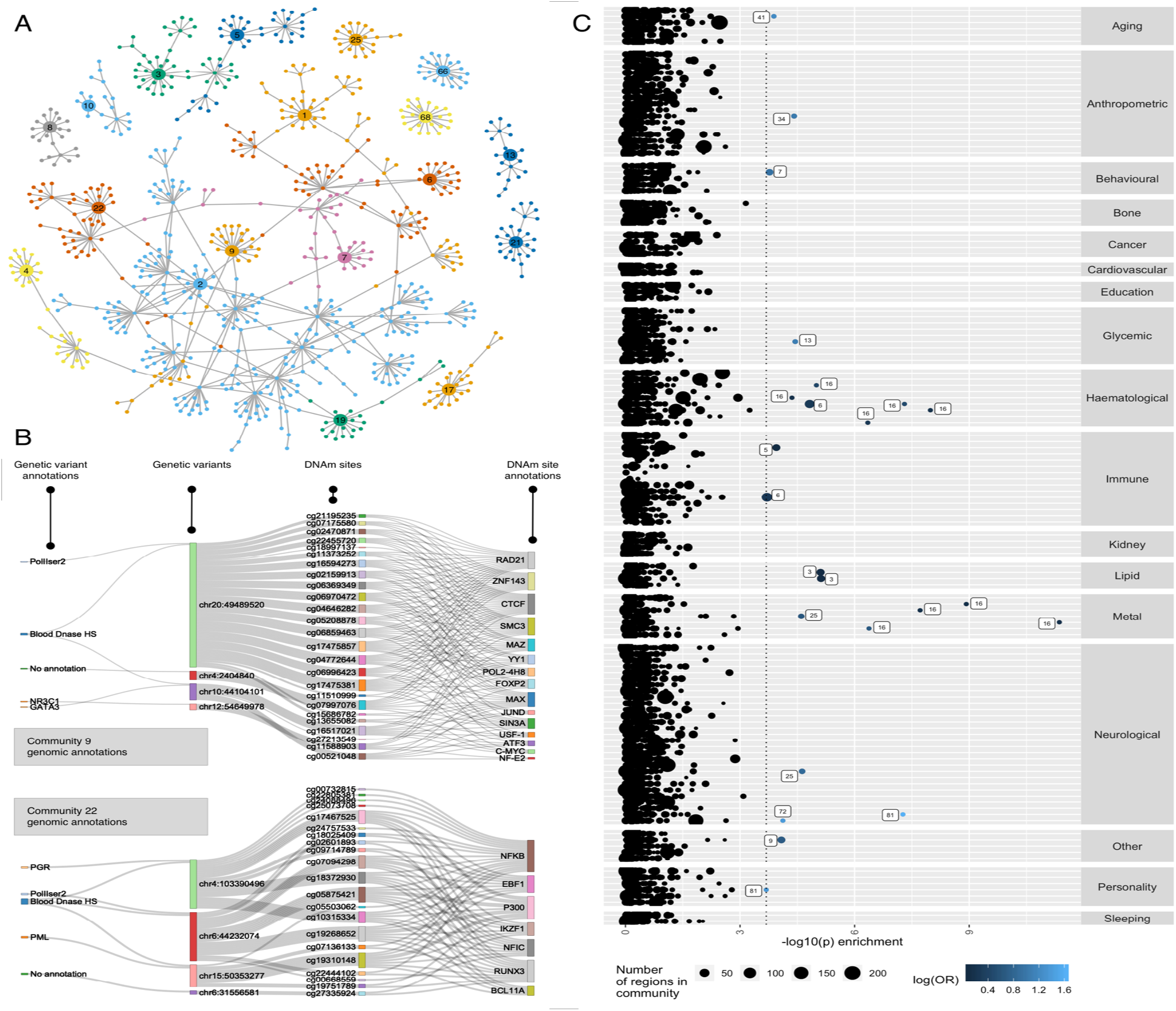
Communities constructed from *trans-*mQTL. **a) A network depicting all communities in which there were twenty or more sites**. Random walks were used to generate communities (colours), so occasionally a DNA site connects different communities. **b) The relationship between genomic annotations, mQTL and communities**. Communities 9 and 22 are comprised of DNAm sites that are related through shared genetic factors. The sankey plots show the genomic annotations for the genetic variants (left) and for the DNAm sites (right). The DNAm sites comprising these communities are enriched for TFBS related to the cohesin complex and NFkB, respectively. **c) Enrichment of GWA traits among community SNPs**. The genomic loci for each of the 56 largest communities were tested for enrichment of low p-values in 133 complex trait GWAS (y-axis). The x-axis depicts the -log10 p-value for enrichment, with the 5% FDR shown by the vertical dotted line. Enrichments were particularly strong for blood related phenotypes (including circulating metal levels).

We hypothesised that *cis* sites were causally influencing multiple *trans* sites within their communities (i.e. a causal chain of mQTL to DNAm at a *cis* site to DNAm at a *trans* site). We evaluated whether the estimated causal effect (obtained from the *trans-*mQTL effect divided by the *cis-*mQTL effect i.e. the Wald ratio) of the *cis* site on the *trans* site was consistent with the observational correlation between the *cis-* and *trans-site*. While there was an association, the relationship was weak (r=0.096, p=1.73e-6, **Figure S30**), indicating that changes in *cis* sites causing changes in *trans* sites is likely not the predominant mechanism. We did observe that the *cis-trans* DNAm levels were more strongly correlated than we would expect by chance (**Figure S31**), which supports the notion that they are jointly regulated without generally being causally related.

To gain functional insights into these communities, we evaluated if DNAm sites within communities were enriched for regulatory annotations and/or gene ontologies (**Table S14-S17, Figure S32-33**). Multiple communities showed enrichments (FDR P <0.001); for example community 9 sites were strongly enriched for TFBS annotations relating to the cohesin complex in multiple cell types, community 22 sites were enriched for NFKB and EBF1 in B lymphocytes and community 76 sites were enriched for EZH2 and SUZ12 and bivalent promotor and repressed polycomb states (**Figure 3B**). Community 2 (comprising 253 sites) was enriched for active enhancer state in 3 cell types and for lymphocyte activation (GO:0046649 FDR p = 0.016) and multiple KEGG pathways including the JAK-STAT signalling pathway (I04630: FDR p=8.53e-7) (**Table S16, Table S17**).

Regulatory features within a network may share a set of biological features that are related to complex traits. We performed enrichment analysis to evaluate if the loci tagged by DNAm sites in a community were related to each of 133 complex traits (**Table S18**), accounting for nonrandom genomic properties of the selected loci. Restricting the analysis to only the 56 communities with ten or more sites, we found eleven communities that tagged genomic loci that were enriched for small p-values with 22 complex traits (FDR < 0.05) (**Figure 3C, Table S19**). Blood related phenotypes were overrepresented (11 out of 23 enrichments being related to metal levels or haematological measures, binomial test p-value = 4.2e-5). Amongst the communities enriched for GWAS signals, community 16 was highly associated with iron and haemoglobin traits. Community 9 was associated to plasma cortisol (p = 8.27e-5). Finally, we performed enrichment analysis on 36 blood cell count traits^35^ and found enrichments for two communities. Community 16 was enriched for hematocrit (p=4.34e-10) and hemoglobin concentration (p=1.99e-8) and community 5 was enriched for reticulocyte traits (p=1.67e-6) (**Figure S34**). The enrichments found for these DNAm communities indicate that a potentially valuable utility of mapping *trans-*mQTL is to indicate how distal regions of the genome are functionally related independent from cellular composition.

### mQTL can be used to identify shared genetic influences with disease

The majority of GWA loci map to non-coding regions^36^ and *cis* mQTL are enriched amongst GWA^17,37,38^. Here we investigated the value of the large number of mQTL especially *trans* mQTL to annotate functional consequences of GWA loci. We first tested genome-wide enrichment of GWAS associations (SNPs at p < 5e-8 for a given complex trait) amongst mQTL SNPs, performing separate analysis for mQTL acting in *cis, cis* and *trans* and *trans*. We utilized genome-wide summary statistics for 37 phenotypes related to 11 disease/trait categories with 41 publicly available GWAS datasets (**Table S20**). After accounting for non-random genomic distribution of mQTL^39^ and multiple testing, we identified enrichments for 35% of the complex traits (**Figure S35, Table S20, Supplementary Information**) mainly for studies with a larger number of GWA signals. The *cis+trans* mQTL were most strongly enriched for low p-values across multiple traits. Six phenotypes across 4 disease categories were associated with *cis* mQTL, nine phenotypes across 5 disease categories were associated with *cis+trans* mQTL.

Inflammatory bowel disease and Crohn’s disease were associated with both sets. Height was associated across all three categories of mQTL but interestingly was depleted for mQTL in the *trans only* group (OR=0.354, p=7.31e-8). The distribution of enrichment effect estimates (ORs) of *trans* mQTL was substantially closer to the null or in depletion when compared to mQTL that included *cis* effects (**Figure 2E**). These enrichments correspond to the results reported earlier, in which *trans-SNPs* were typically found in intergenic regions, which are known to be depleted for complex trait loci^40^.

Though the mQTL discovery pipeline adjusted for predicted cell types^41,42^ and non-genetic DNAm PCs, there is a possibility that residual cell-type heterogeneity remains. We performed another set of GWAS enrichment analysis, this time using 36 blood cell traits^35^, and found enrichments. These were strongest amongst *cis+trans* mQTL, as seen in the previous enrichments (**Figure S36**). Interrogating this further, we found that for 98.9-100% of the mQTL, mQTL SNPs explained more variation in DNAm than they explain variation in blood cell counts suggesting a causal chain of mQTL to blood trait^43^. Alternatively, a systematic measurement error difference could explain these observations, where DNAm captures blood cell counts more accurately than conventional measures.

The enrichments suggest that overlaps are not due to chance which motivated us to a much more in-depth analysis on a much larger number of traits/diseases. We searched for instances of DNAm sites sharing the same genetic factors against each of 116 complex traits and diseases, and initially found 23,139 instances of an mQTL strongly associating with a complex trait (**Figure 4**). To evaluate the extent to which these were due to shared genetic factors (and not, for example, LD between independent causal variants), we performed genetic colocalization analysis^44^ (**Table S18, Table S21**). Excluding genetic variants in the *MHC* region, we found 1,373 putative examples in which at least one DNAm site putatively shared a genetic factor with at least one of 71 traits (including 19 diseases). Those DNAm sites that had a shared genetic factor with a trait were 6.9 times more likely to be present in a community compared to any other DNAm site with a known mQTL (Fisher’s exact test 95% CI 4.8-9.7, p =9.2e-19). Next, we evaluated how often the DNAm site that colocalised with a known GWAS hit was the closest DNAm site to the lead GWAS variant by physical distance. Notably, in only 18.1% of the cases where a GWAS signal and a DNAm site colocalised, was that DNAm site the closest DNAm site to the signal. This finding is similar to results found for gene expression^45^, but the converse has been found for protein levels^46^.

**Figure 4:**
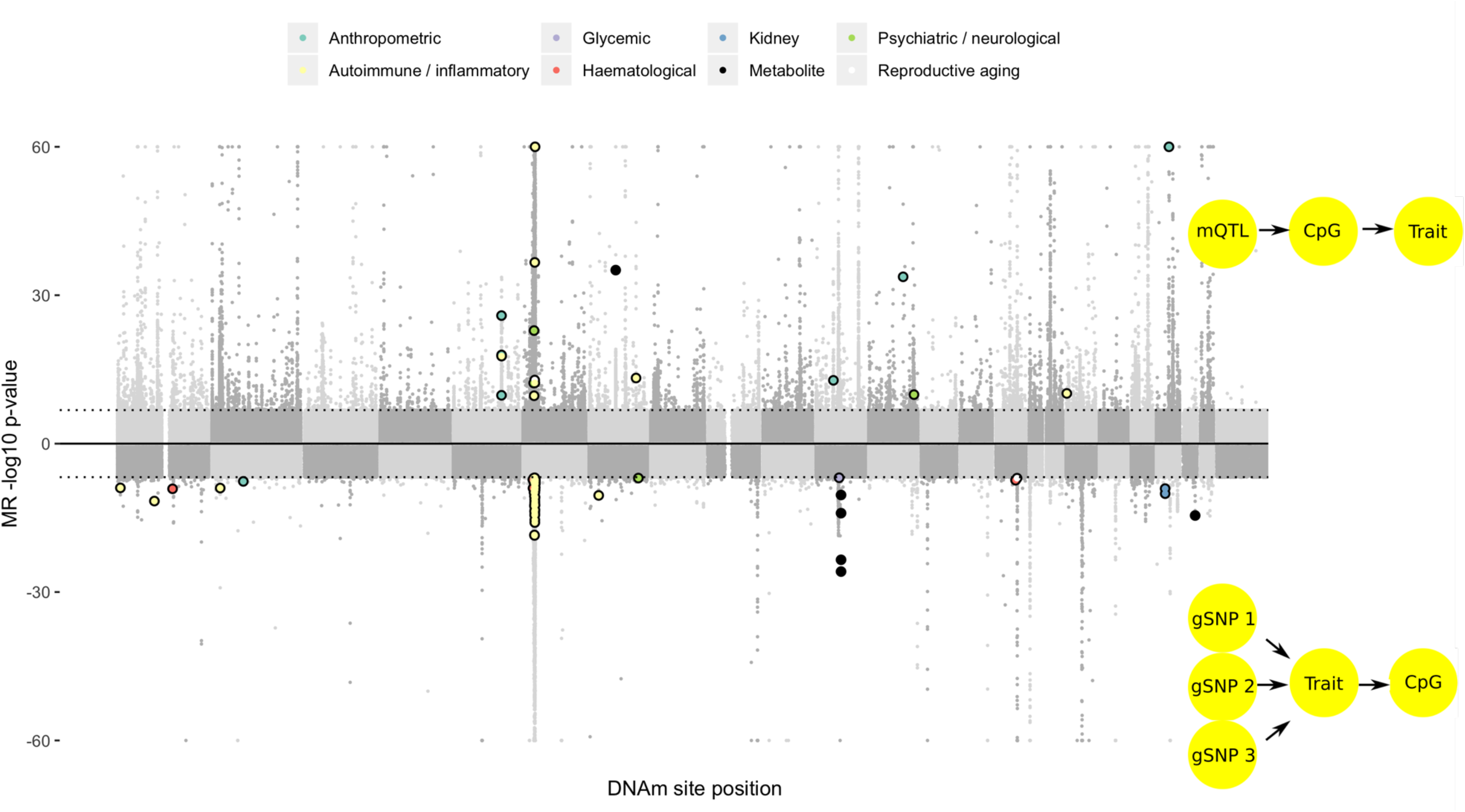
Identifying putative causal relationships between sites and traits using bidirectional MR. Aggregated results from a systematic bi-directional MR analysis between DNAm sites and 116 complex traits. The top plot depicts results from tests of DNAm sites colocalising with complex traits. The light grey points represent MR estimates that either did not surpass multiple testing, or shared small p-values at both the DNAm site and complex trait but had weak evidence of colocalisation. Bold, coloured points are those that showed strong evidence for colocalisation (H4 > 0.8). The bottom plot shows the –log10 p-values from MR analysis of risk factor or genetic liability of disease on DNAm levels. Extensive follow up was performed on DNAm site-trait pairs with putative associations, and those that pass filters are plotted in bold and colored according to the trait category. A substantial number of MR results in both directions exhibited very strong effects but failed to withstand sensitivity analyses.

It has previously been difficult to conclude whether genetic colocalisation between DNAm and complex traits indicates a) a causal relationship where the DNAm level is on the pathway from genetic variant to trait (vertical pleiotropy) or b) a non-causal relationship where the variant influences the trait and DNAm independently through different pathways (horizontal pleiotropy)^47^. In Mendelian randomisation (MR) it is reasoned that under a causal model, multiple independent genetic variants influencing DNAm should exhibit consistent causal effects on the complex trait^48^. Amongst the putative colocalising signals, 440 (32%) involved a DNAm site that had at least one other independent mQTL. To test if there was a general trend of DNAm sites causally influencing a trait, we evaluated if the MR effect estimate based on the colocalising signals were consistent with those obtained based on the secondary signals. There were substantially more large genetic effects of the secondary mQTL on respective traits than expected by chance (70 with p < 0.05, binomial test p = 2.4e-16). However only 41 (59%) of these had effect estimates in the same direction as the primary colocalising variant, which is not substantially better than chance (binomial test p = 0.19). Twelve of the 41 mQTL were located in the *HLA* region. Of the remaining mQTL, 27 were associated with anthropometric (*ESR1* and birth weight), immune response (*IRF5* and systemic lupus erythematosus) and lipid traits (*TBL2* and triglycerides). We then performed systematic colocalization analysis of all mQTL against 36 blood cell traits^35^. Here we discovered 94,738 instances of a DNAm site and a blood cell trait sharing a causal variant. In 28,138 instances the colocalising DNAm site had an independent secondary mQTL, and with these associations we again tested for a general trend of DNAm sites causally influencing the blood trait. The association between primary and secondary signals was very weak (R^2^ = 0.008), suggesting that the general causal model is not supported. Together, these results indicate that those blood measured DNAm sites that have shared genetic factors with traits cannot be typically thought of as mediating the genetic association to the trait (**Figure S37-S38, Table S22**). Instead, if DNAm is a coregulatory phenomenon then the colocalising signals between DNAm sites and complex traits may be due to a common cause, for example genetic variants primarily acting on TF binding^8,49,50^.

### The influence of traits on DNAm variation

Previous studies have not been adequately powered to estimate the causal influences of complex traits on DNAm variation through MR, as the sample size of the outcome variable (DNAm) is a predominant factor in statistical power^44,51^. We systematically analysed 109 traits for causal effects on DNAm using two-sample MR^52,53^, where each trait was instrumented using SNPs obtained from their respective previously published GWAS (**Supplemental Note 2, Table S18**). Included amongst the traits were 35 disease traits, which when used as exposure variables in MR must be interpreted in terms of the influence of liability rather than presence/absence of disease. The sample size used to estimate SNP effects in DNAm was up to 27,750 (**Figure 4**).

We initially identified 4785 associations where risk factors or genetic liability to disease influences DNAm levels (multiple testing threshold p < 1.4e-7). However, MR analysis on omic variables can lead to false positives due to violations in assumptions. We developed a filtering process involving a novel causal inference method to help protect against these invalid associations (**Supplementary Information, Supplementary Note 2, Figure S39**). This left 85 associations (involving 84 DNAm sites) in which DNAm sites were putatively influenced by 13 traits (nine risk factors or four diseases) (**Table S23**). Further filtering that would exclude traits that were predominantly instrumented by variants in the *HLA* region or driven by one SNP would reduce the total number of associations substantially from 84 to 19. We replicated five associations for triglycerides influencing DNAm sites near *CPTA1* and *ABCG1^54^* and found associations for transferrin saturation/iron influencing DNAm sites near *HFE*.

We next evaluated if there was evidence for small, widespread changes in DNAm levels in response to complex trait variation, by calculating the genomic control inflation factor (GC_in_) for the p-values obtained from the MR analyses of each trait against all DNAm sites. Five traits (fasting glucose, age at menarche, cigarettes smoked per day, immunoglobulin G index levels, serum creatinine), showed GC_in_ values above 1.05 (**Figure S40**). A high GC_in_ value can be the result of the trait that has an influence on a few sites or has a widespread effect on DNAm. GC_in_ calculations were performed at each chromosome singly for each trait (**Figure S41**) and in a leave-one-chromosome-out analysis (**Figure S42**). The GC_in_ remained consistent (except for immunoglobulin G index levels), indicating that the traits have small but widespread influences on DNAm levels across the genome.

While most of the traits (n=105, 96%) tested did not appear to induce genome-wide enrichment this does not rule out the possibility of them having many localised small effects. For example, the smallest MR p-value for the analysis of body mass index on DNAm levels was 2.27e-6, which did not withstand genome-wide multiple testing correction, and GC_in_ was 0.95. However, restricting GC_in_ to 187 sites known to associate with body mass index from previous epigenome-wide association studies (EWAS)^20^ indicated a strong enrichment of low p-values (median GC_in_ = 3.95). A similar pattern was found for triglycerides, in which genome-wide median GC_in_ = 0.94 but the 10 sites known to associate with triglycerides from previous EWAS^55^ had an MR p-value of 8.3e-70 (Fisher’s combined probability test). These results indicate that traits causally influencing DNAm levels in blood is the most likely mechanism that gives rise to these EWAS hits. It also indicates that the general finding that there were very few filtered putative causal effects of risk factors or genetic liability to disease on DNAm could be due to true positives being generally very small, even to the extent that our sample size of up to 27,750 individuals was insufficient to find them.

### DNAm sites influenced by genetic variation are under selection

Natural selection has modified the allele frequency of complex trait associated variants through their beneficial or deleterious effects on fitness^56,57,58,59^. Here we investigate whether mQTL SNPs are frequent targets of natural selection utilizing selection scores acting through different timescales and mechanisms to each SNP in 1000G: a population differentiation method (global F_st_), several haplotype-based methods (integrated haplotype score (iHS), Cross Population Extended Haplotype Homozygosity (XPEHH) and the singleton density score (SDS) (**Table S24, Supplementary Information**).

We then tested whether there is enrichment of mQTL associations (Bonferroni adjusted p <0.01) among SNPs that show evidence of positive selection for each metric while controlling for non-random genomic distribution^39^ (excluding two regions (*HLA* and *LCT*) known to be under high selective pressure). We found enrichments of positive selection signatures among SNPs with *cis only* (*F*_st_: p=7.87e-23, OR=1.31, SDS: p=4.43e-10, OR=1.42) and *cis+trans* (*F*_st_: p=7.1e-21, OR=1.35, SDS: p=4.35e-11, OR=1.53, XPEHH (CEU vs CHB): p=7.7e-7, OR=1.53) associations (**Figure 2F, Table S25**). The strong enrichments for *cis+trans* (n=107-1585) and *cis only* (n=1186-4980) indicating that positive selection is most likely to operate on *cis* acting variants. However, there is less power to detect these enrichments for *trans only* SNPs (n=14-102).

We next examined whether there was a relationship between the mQTL effect sizes (allele frequency adjusted) and the selection scores as a proxy for the estimated strength of selection. Using a linear model for each of the selection metrics (accounting for the number of proxies, distance to TSS, CpG and GC frequency), we found that the strongest mQTL effect size was positively associated with F_st_ (p<1.1e-05) but not with recent changes in allele frequency (measured by SDS) with consistent directions across the mQTL categories (*cis only, cis+trans* and *trans only*) (**Figure S43**). These results may indicate that DNA sites might either the primary target of selection or the mQTL SNP have pleiotropic effects on fitness^60^.

Enrichment of F_st_ amongst mQTL could also be due to negative selection. Evidence for negative selection can be inferred from the strong negative relationship between mQTL SNP effect size and MAF (difference in mQTL SNP effect size=-0.56, p=2.2e-308, **Figure S43**). To confirm that this relationship is not an artefact of having defined the SNP effect via the maximum effect each SNP has on any DNAm site, we developed a novel method (**Supplementary Information, Figure S44**) to quantify the relationship for the strongest acting SNPs at a given frequency, allowing for a majority of unselected SNPs. SNPs with a higher frequency have a smaller average effect (S=0.4, CI 0.325-0.475), where S=0 corresponds to no selection and S=1 corresponds to strong negative selection. We found similar relationships across the mQTL categories (*cis only, cis+trans* and *trans only*) (**Figure S45**) though there was insufficient power to quantify selection for *trans only* SNPs. These results can be interpreted that predominantly genetic regions that regulate DNAm are under negative or balancing selection^60,61^ and thus, retain the ancestral DNAm structure. However, a minority of regions containing DNAm sites have experienced positive selection.

Alleles showing evidence of selection are likely to be biologically meaningful^62^. To investigate whether genetic variants underlying DNAm implicated in selection are linked to diseases/traits, we examined whether GWAS-associated variants from 42 datasets across 11 disease categories were enriched for *cis* mQTL SNPs overlapping extreme SDS scores. After accounting for non-random genomic distribution^39^, we found that GWAS-associated variants from 19/42 traits were overlapping with at least one *cis* mQTL SNP with extreme SDS. We found an enrichment of mQTL SNPs overlapping extreme SDS scores (p<2.6e-3) among variants associated with five traits including extreme height (OR=17.2, p=1.08e-7), Crohn’s disease (OR=11.3, p=4.42e-5), height (OR=1.99, p=6.76e-5), schizophrenia (OR=5.28, p=1.21e-3) and cardiovascular disease (OR=9.85, p=1.67e-3) (**Table S26**). A comparison showed that the genetic variance for cardiovascular disease associated mQTL or height associated mQTL with extreme SDS was higher when compared to all trait associated SNPs suggesting that these loci have been modified by disease pressure (**Figure S46**). To summarize, our results provide the first evidence that selection may have shaped the landscape of DNAm values across the genome and is associated with adaptation on highly polygenic (height) or fitness related (cardiovascular) traits.

## Implications

A map of hundreds of thousands of genetic associations has enabled novel biological insights related to DNAm variation. Using a rigorous analytical framework enabled us to minimise heterogeneity and expand sample sizes for large omic data. This revealed a genetic architecture of DNAm that is polygenic, as with many traits traditionally considered ‘complex’. Given the diverse ranges of age, gender proportions and geographical origins between the cohorts in this analysis, the minimal extent of heterogeneity across datasets indicates that genetic effects on DNAm are relatively stable across contexts. We show that *cis* and *trans* mQTL operate through distinct mechanisms, as their genomic properties are distinct. A driver of long-range associations may be co-regulated through TF binding and nuclear organisation.

Though we found substantial sharing of genetic signals between DNAm sites and complex traits, we were able to demonstrate that this was not predominantly due to DNAm variation being on the causal path from genotype to phenotype. These findings have several implications. First, we anticipate that some previously reported EWAS associations are likely due to reverse causation e.g. the risk factor or genetic liability to disease state itself alters DNAm and not vice versa, or confounding. Second, having found there are strong negative and positive selection pressures acting on mQTL, this may be explained through selection acting on complex traits first. Third, the genetic effects on DNAm that overlap with complex traits likely primarily influence other regulatory factors which in turn influence complex traits and DNAm through diverging pathways. Fourth, if the path from genotype to complex traits is non-linear, for example involving the statistical interactions between different regulatory features^16^, then our results indicate that large individual-level multi-omic datasets will be required to dissect such mechanisms.

The microarray technology used in the majority of cohorts included in this study limited us to analyse only 2% of sites across the genome^63^, which are biased to promoters and strongly underrepresented regulatory elements. To explore the impact of expanding the coverage of arrays, we calculated the linear relationship between the median number of probes by gene on the 450k array and the median number of *cis* and *trans* mQTL. For each probe, we found an increase of 0.76 *cis* mQTL (p<9.03e-16) and 0.05 *trans* mQTL (p<1.47e-05) in the median (**Figure S47**). A similar increase was seen in non-genic regions. EPIC arrays^63^ and promising low-cost sequencing technologies^64^ offer an opportunity to make detailed interrogations of enhancer and other regulatory regions, which may be more fruitful in finding causal relationships with complex disease.

The coverage of the mQTL search in this study was limited by the computational necessity of a multiple stage study design (**Figure S48**). Those mQTL that we discovered with r^2^ less than 1% are likely a small fraction of all the mQTL in this category expected to exist (**Figure S49**). Across these DNAm sites, and within the range of mQTL detected in our study (r^2^ > 0.22%) we estimate that there are twice as many *cis* mQTL and 22.5 times more *trans* mQTL yet to discover (**Figure S49**). This would likely not explain all estimated heritability, indicating that a substantial set of the heritability is due to causal variants with smaller effects than those detectable in our study.

Overall our data and results have resulted in the most comprehensive atlas of genetic effects to date. We expect that this atlas will be of use to the scientific community for studies of genome regulation and contribute to the development of second generation EWAS.

## Data Availability

A database of our results is available as a resource to the community at http://mqtldb.godmc.org.uk. The individual level genotype and DNAm data are available by request from each individual study or can be downloaded from Gene Expression Omnibus as described in Supplemental Note 1. As the consents for most studies require the data to be under managed access, the individual level genotype and DNAm data are not available from a public repository unless stated in Supplemental Note 1.

## Contributions

**Project management:** G.S., J.L.M

**Designed individual studies and contributed data:**

A.A.C., A.Cas., A.D.H., A.G.U, A.Me., A.Mu., A.M.M., B.B., B.T.H., C.H., C.L.R., C.P., C.Sa., C.Sh., C.Sö., D.A.L., D.v.H., D.I.B., D.T., E.A.N., E.B.B., E.J.C.d.G, E.M., F.G., F.R., G.E.D, G.H.K., G.P., G.W.M., H.R.E., H.T., H.Z., I.J.D., J.F.F., J.H.V., J.J.C., J.Ka., J.L., J.M., J.M.S., J.M.V., J.v.M., J.R., J.R.B.P., J.R.G., J.Sh., J.T.B., J.W., J.W.H., K.K.O., K.L.E., K.R., L.A., L.C.S., L.M., M.A.I., M.Bee., M.Bu., M.E.A.R., M.H.v.IJ., M.Ke., M.O., N.C., N.G.M., N.J.W., N.R.W., P.E.S., P.Mo., P.M.V., R.H., R.P., S.L., T.D.S., T.E., T.E.M., T.I.A.S, T.P., T.T., V.W.V.J., W.K., Z.P.

**Generated and/or quality-controlled data:** A.A.K., A.I., A.S., C.S.M., H.R.E., J.L.M., K.B., K.M.H., N.K., S.M.R., T.H., R.M.W., W.L.M.

**Designed new statistical or bioinformatics tools:**

G.H., J.L.M., M.Su., T.R.G., V.I.

**Analysed the data and/or provided critical interpretation of results:**

A.D.B, A.Car., A.D., A.F.M., A.K., B.T.H., C.B., C.H., C.L.R., C.R.A., C.Sor., C.V., C.X., C.W., D.A., D.C., D.J.L., D.L.C., D.M., E.C.M., E.G., E.H., E.M., F.C.M., F.I.R., F.R.D., G.B., G.C., G.D.S., G.H., G.H.K., G.M., G.W., I.Y., J.C.F., J.v.D., J.J.H., J.Ka., J.Kl., J.L.M., J.M., J.Su., J.T.B., K.B., K.v.E., K.F.D., K.S., L.C.S., M.Ber., M.Bu., M.H.v.IJ., M.G., M.Ku., M.L., M.Sm., M.Su., N.K., P.Me., P.Ma., P.M.V., R.E.M., R.G., R.L., R.Z., S.B., S.G., S.K., T.C., T.G., T.G.R., T.I.A.S., T.L., T.R.G., Y.A., Y.Z., V.I., V.S.

**Designed and/or managed the study:** B.T.H., C.B., C.L.R., J.M., J.T.B., T.R.G.

**Wrote the manuscript:** A.D.B., B.T.H., C.B., C.L.R., D.J.L., E.C.M, E.H., G.D.S., G.H., J.C.F., J.Kl., J.L.M., J.M., J.T.B., K.B., K.F.D., M.Su., P.M.V., R.L., T.G.R., T.R.G., V.I.

